# Mobility-informed metapopulation models predict the spatio-temporal spread of respiratory epidemics across scales

**DOI:** 10.1101/2025.06.26.25330297

**Authors:** Aakash Pandey, Lu Zhong, Lior Rennert

## Abstract

Understanding the spatiotemporal dynamics of infectious disease spread is critical for anticipating epidemic trajectories and guiding public health responses. Accurate forecasts of where and when outbreaks are likely to emerge can support efficient resource allocation, particularly during the early stages of epidemics when surveillance data are limited. In this study, we used empirical human mobility data derived from county-level commuting and air traffic flows, and a theoretical mobility model (the radiation model) to study the relative order of epidemic onset across spatial scales. These mobility models were incorporated into a metapopulation framework to predict the spread of three major respiratory pathogens: COVID-19, seasonal influenza, and respiratory syncytial virus (RSV). We applied this framework to county-level transmission within South Carolina and state-level introductions across the United States. In both empirical and theoretical mobility scenarios, we found that effective distance, a network-based measure of mobility-informed proximity, reliably predicts the relative timing of epidemic onset. These results demonstrate that mobility-informed metapopulation models can capture consistent spatiotemporal patterns across disease systems and spatial scales, even in the absence of detailed epidemiological parameters. This highlights their potential as scalable, data-efficient tools for outbreak forecasting and early public health planning.

## Introduction

Accurate predictions of when and where infectious diseases are likely to spread next are critical for improving the effectiveness of public health interventions. During the early stages of an epidemic, epidemiological data are sparse, and uncertainty is high [1,2]. Despite this, health authorities must often make rapid decisions about the allocation of limited resources, including personnel, diagnostic tools, pharmaceutical and non-pharmaceutical interventions. Strategic deployment of these constrained resources is essential to reduce transmission, mitigate morbidity and mortality, and prevent waste [3,4]. Mathematical modeling can serve as a vital tool in this decision-making process, offering insights into transmission dynamics and guiding intervention strategies [5,6].

Metapopulation models are widely used in the study of the spatio-temporal dynamics of infectious disease spread [7–10]. Central to these models is the incorporation of human mobility, which governs the spatial spread and temporal progression of outbreaks [11–14]. Metapopulation models parametrized with empirical data or with theoretical models can be used in forecasting disease arrival times and transmission trajectories. For instance, the Global Epidemic and Mobility (GLEaM) model, which integrated airline travel data and commuting patterns, was used to predict the spread of the 2009 H1N1 influenza pandemic across international and subnational scales [15–17]. Similar studies have been carried out to reproduce the spatial diffusion pattern of diseases like HIV [18] and SARS 2001[19,20]. More recently, data-informed models have been central to understanding COVID-19 dynamics. Kraemer et al. (2021) leveraged mobile phone-derived human mobility data to track and predict the spread of SARS-CoV-2 lineage B.1.1.7 in the UK [21], while Pei et al. (2021) applied commuting data to model the spatial propagation and differential impacts of intervention timing on COVID-19 within the United States [22]. Similarly, Hamada et al. (2020) found a strong correlation between mobility patterns and COVID-19 early case growth rates at the US county level [23]. These studies demonstrate that mobility-based models can yield probabilistic forecasts of disease spread, including the relative order and timing of outbreak emergence across regions, and can assess the impact of different interventions in reducing the transmission.

Respiratory infectious diseases, including COVID-19, seasonal influenza, and respiratory syncytial virus (RSV), remain a substantial public health burden within the United States and across South Carolina [24]. For example, based on the recent FluSurv-NET data, the cumulative hospitalization rate for the 2024/2025 influenza season was the highest observed since the 2010-2011 season [25]. Likewise, during the peak time in the 2024/2025 flu season, about 13% of all emergency department visits were due to influenza-related infections, while RSV accounted for about 1.35% of all ED visits during its peak time in South Carolina [26]. Similarly, between March 2020 and December 2023, there were more than 360,000 hospitalizations due to COVID-19 alone [27]. The capacity to monitor and anticipate the geographic spread of such pathogens is critical for effective planning and resource allocation. This need is particularly acute in South Carolina due to pronounced disparities in healthcare infrastructure between rural and urban regions [28]. The ability to estimate both the likelihood and relative timing of disease arrival across the state is essential for informing strategic planning and mitigating the disproportionate impact on vulnerable populations across the state.

In this study, we develop a mobility-informed metapopulation framework to investigate the spatio-temporal dynamics of respiratory disease spread in the U.S at the state level, and within South Carolina counties. Specifically, we utilize county-level commuting data to estimate the relative order of COVID-19 arrival during the first epidemic wave, as well as the seasonal spread of influenza and respiratory syncytial virus (RSV) across multiple years within South Carolina. We address the question: Can readily available mobility datasets, such as county-to-county commuting flows and airline traffic volumes, be used to predict the relative timing of disease arrival and epidemic onset at both state and sub-state (county) levels? We demonstrate that a simple mobility data-informed network-based metric, effective distance, can be used to reconstruct the geographic progression of infectious disease spread with notable accuracy. Furthermore, we show that theoretical mobility models, such as the radiation model, can serve as viable alternatives and yield comparable spatial predictions. Our findings underscore the potential of mobility-driven models as low-data, high-utility tools for anticipating epidemic trajectories and supporting proactive public health responses.

## Methods

### Data sources

COVID-19 case data at the state level across the United States was taken from the cumulative data that is publicly available on the New York Times GitHub repository [29]. The state-level influenza data were obtained from the CDC Flu View website [30]. The state-level mobility data were obtained from the U.S. Department of Transportation consumer airfare report, which has the number of passengers between the top 1000 contiguous city pairs markets that average at least 10 passengers per day [31]. Weekly COVID-19, RSV, and influenza inpatient hospitalization data at the county level were obtained from the South Carolina Revenue and Fiscal Affairs Office for years ranging from 2019 to 2023. County-level population and commuting data were obtained from the US Census Bureau[32].

### Mobility model and effective distance

The effective distance between counties was calculated using a theoretical mobility model and commuting data. For the theoretical model, the mobility flux was estimated using the radiation model [33]. The effective distance between counties *i* and *j* is defined as [7],

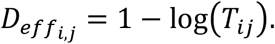

*T*_*ij*_ represents the average mobility flux for the county *i* to county *j* and is calculated as

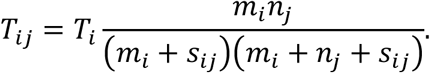

*T*_*i*_ represents the total number of commuters originating from the county *i. m*_*j*_ and *n*_*j*_ represent the population sizes of the counties *i* and *j*, respectively. Similarly, *s*_*ij*_ is the total population in the circle of radius *r*_*ij*_ centered at *i* but excludes the *m*_*i*_ and *n*_*j*_.

### Mobility data and effective distance

To quantify inter-county connectivity within South Carolina based on human mobility, we derived a directed, weighted commuting network using the 2022 U.S. Census Bureau commuting flow data [32]. The dataset provided estimates of daily commuting volume between county pairs, with each observation denoting the number of individuals commuting from an ‘origin’ county *i* to a ‘destination’ county *j* (denoted *F*_*ij*_). The data was converted into an *N* × *N* origin-destination matrix, where *N* is the number of counties, and each element *F*_*ij*_ represents the number of commuters from the county *i* to county *j*. Missing values in the matrix were set to zero, assuming no commuter flow. To construct a stochastic transition matrix *P*, we normalized each row of the adjacency matrix by the total outflow from the origin county as

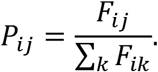

Elements corresponding to commuting within the same counties were also set to zero. The resulting origin-destination matrix was used to construct a directed, weighted graph *G* using the *igraph* package in R (v1.5.1) [34]. The weights of the edges were assigned as − log(*P*_*ij*_). To assess the minimum effective distances between counties, we computed all-pairs shortest paths on the full graph *G*, treating the effective distances as edge weights and using a Dijkstra-based algorithm in *igraph*. The resulting matrix *D*_*eff*_ captures the shortest effective-path length from each county to every other county, accounting for direct and indirect commuter linkages.

### Estimating epidemic onset time

Surveillance and case reporting data are often incomplete and sensitive to various underlying surveillance structures and public health resources. This is exacerbated at a fine-scale geographic locations and in rural areas that lack robust public health infrastructures. In these scenarios, case reporting at or near epidemic peak can be more reliable to infer overall trends in epidemic dynamics. Hence, to estimate the temporal onset of local epidemics across counties, we used a retrospective modeling approach based on the analytical solution of the classic susceptible–infectious–recovered (SIR) model. This method infers the likely time of epidemic seeding by leveraging observed peak infection times and an assumption of exponential early growth constrained by the basic reproduction number (*R*_0_).

Weekly county-level inpatient data were first smoothed to reduce noise using a LOESS (locally estimated scatterplot smoothing) function. The maximum of the smoothed curve for each county was identified, denoting both the date and magnitude of peak hospitalization. We assumed that inpatient hospitalization data represented a fixed proportion of the true infectious prevalence and scaled the smoothed peak values by the respective hospitalization factor for each disease. For example, although the case-to-hospitalization ratio varies during different waves of COVID-19 [35], early estimates before 2021 show that about 5% of estimated infections were hospitalized [36]. Hence, we scale the peak weekly COVID-19 hospitalization by 20x to estimate the peak weekly infections at the county level. The scaled peak prevalence was used to infer the county-level basic reproduction number (*R*_0*i*_) assuming a simple SIR model dynamics using the following analytical approximation [37],

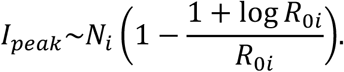

Here, *I*_*peak*_ is the scaled peak prevalence, *N*_*i*_ is the susceptible population size of each county. Given the estimated *R*_0*i*_, we then calculated the time elapsed from epidemic onset to peak prevalence under deterministic SIR dynamics as:

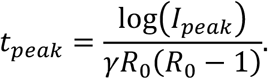

*γ* represents the recovery time. The epidemic onset time was then calculated by subtracting the *t*_*peak*_ from the observed peak dates for each county. We similarly calculated onset dates for influenza and RSV at the county level. Although the case-to-hospitalization ratio varies by age and seasons for both diseases [38,39], for simplicity, we assume the ratio to be around 85 [40].

### Estimating spatial spread from effective distance and epidemic onset times

To infer likely spatial origins of epidemic spread, we assessed the correlation between effective distance and estimated onset timing across all counties. For each focal county *i*, we tested the hypothesis that spatial propagation radiated outward from *i* by correlating effective distances from *i* to all other counties (*D*_*ij*_) with their respective onset times. We identified the earliest-seeded county based on estimated onset dates. The onset dates were transformed into numeric values by subtracting the earliest onset week in the dataset, yielding a time lag variable Δ*t*_*j*_ = *t*_*j*_ − min (*t*_*onset*_), where *t*_*j*_ is the onset week in the county *j*. We then computed the Pearson correlation coefficient between effective distance and onset lag as follows:

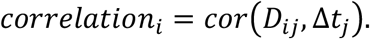

As an alternative approach, correlation was calculated for all counties, which were then ranked by correlation strength, with higher correlations indicating a stronger linear relationship between effective distance and timing that can represent a diffusion-like spread from that focal county [7].

### Metapopulation model

To assess the validity of the modeling framework, we simulated the spatial spread of a hypothetical disease across counties using a discrete-time, stochastic metapopulation SIR model, with inter-county movement defined by the radiation model and commuting data separately. Each county was modeled as a distinct patch with fixed population size, and transmission occurred both locally and via between-county interactions. The epidemic was seeded in a single county (Greenville County), with all other counties initialized with their entire population in the susceptible compartment.

The model progressed over discrete daily timesteps. At each step, within-county transmission was simulated using binomial draws. Cross-county infection was modeled by adding infections due to contact with infectious individuals from other counties, scaled by a transition probability matrix derived from the radiation model and from the commuting data. We ran 1,000 simulations, each starting from the same single-seed initial condition. In each run, the epidemic onset time for a county was recorded as the first day on which the number of infectious individuals exceeded zero. Mean onset times were calculated across simulations for each county. These mean simulated onset times were then correlated with effective distances derived from the radiation-model-based mobility network, as well as commuting network data, to evaluate the predictive relationship between the theoretical effective distance and spatial epidemic timing.

## Results

### Diseases emerge in large counties, followed by gradual diffusion to smaller counties

We observed a consistent pattern in which epidemics emerged earlier in larger, more connected counties, followed by a gradual spread to less populated regions. Specifically, estimated epidemic onset dates were negatively correlated with county population size across all three respiratory pathogens, indicating earlier onset in counties with larger populations. This relationship was strongest for the first wave of COVID-19 (Spearman’s ρ = –0.75, p<0.001; Figure 1c) but was also evident for the 2022–2023 influenza season (ρ = – 0.64, p<0.001; Figure 1d) and the 2022–2023 RSV season (ρ = –0.57, p<0.001; Figure S1). Furthermore, our estimated epidemic onset timings showed moderate to strong correlation with observed hospitalization onset data, with Spearman’s ρ values of 0.64 (p<0.001) for COVID-19 (Figure 1a), 0.73 (p<0.001) for influenza (Figure 1b), and 0.67 (p<0.001) for RSV (Figure S1).

**Figure 1.**
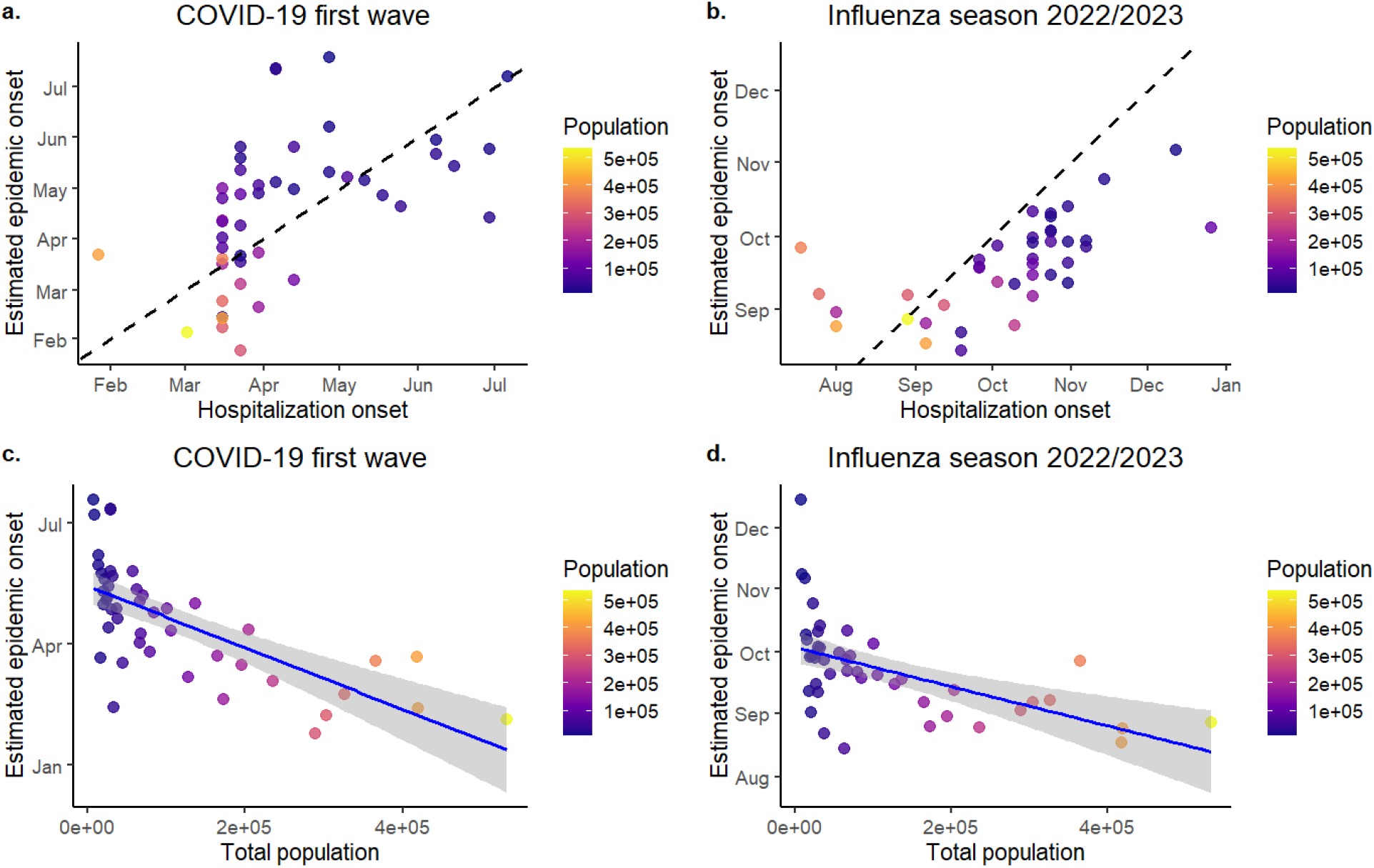
Estimated epidemic onset times for the COVID-19 first wave (left panel) and the influenza 2022/2023 season (right panel). The upper panel (Figures a. and b.) shows comparisons between the estimated onset dates at the county level with observed hospitalization dates (defined as >1 hospitalization). The lower panel (Figures c. and d.) shows correlations between estimated epidemic onset times and population size of each county.

### Effective distance predicts the spatial pattern of epidemic onset

We found that effective distance is a strong predictor of the relative timing of disease spread. In simulations using a metapopulation SIR model with disease introduction in Greenville County, the time to first infection in each county was highly correlated with effective distance from Greenville, both when using the radiation model (Spearman’s ρ = 0.93, p<0.001, Figure 2a) and when using county-level commuting data (ρ = 0.57,p<0.001, Figure S2a). This predictive relationship was also observed in empirical data. Estimated epidemic onset dates during the first wave of COVID-19 were positively correlated with effective distance from Greenville County (ρ = 0.66, p<0.001, using the radiation model; ρ = 0.40, p=0.007, using commuting data). Similar patterns were found for seasonal respiratory viruses, assuming Charleston County as the initial seeding location. For the 2022–2023 influenza season, correlations with effective distance were ρ = 0.69, p<0.001 (radiation model) and ρ = 0.51,p<0.001 (commuting data). For the 2022–2023 RSV season, correlations were ρ = 0.66, p <0.001 (radiation model) and ρ = 0.45,p=0.004 (commuting data).

**Figure 2.**
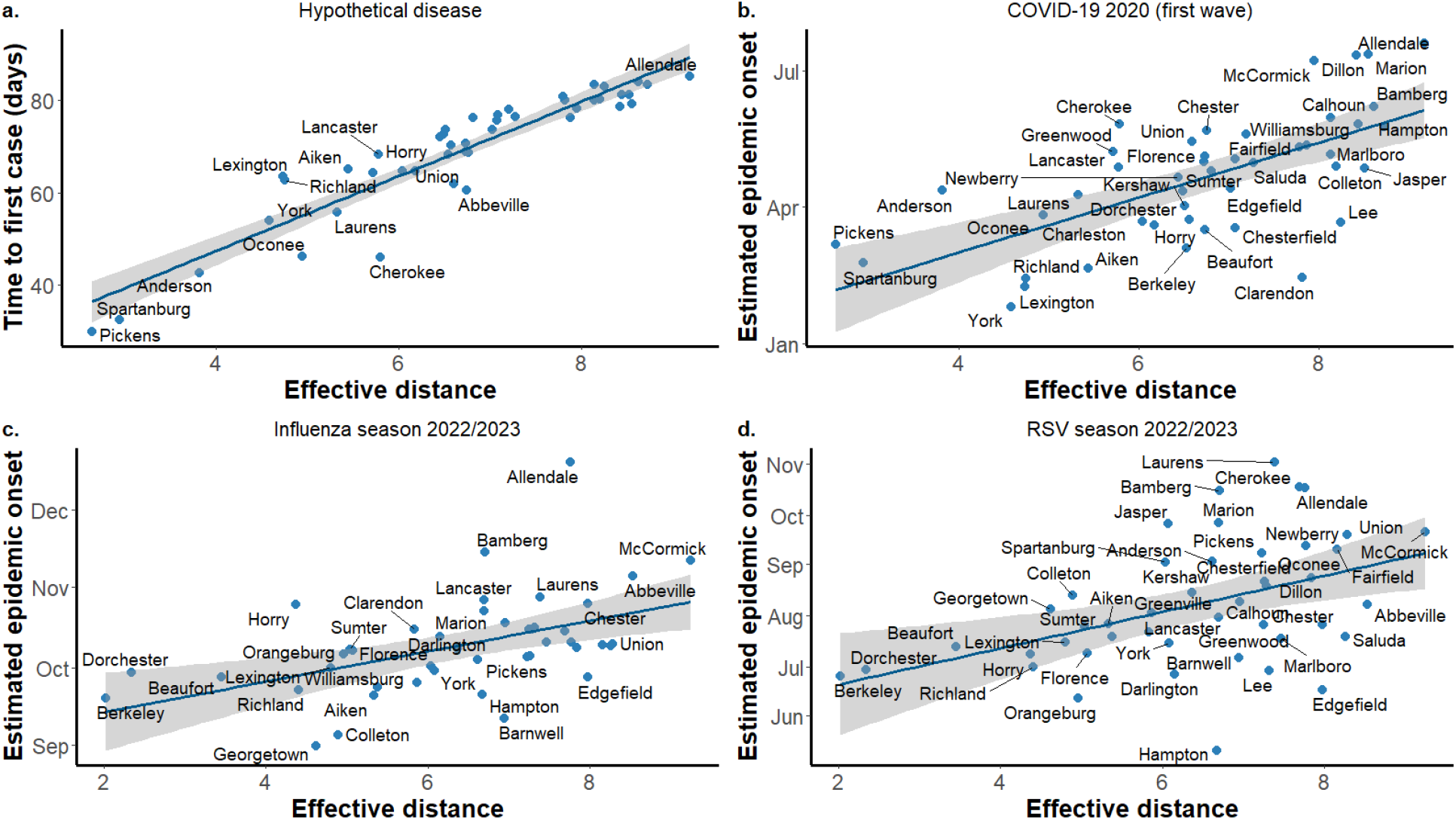
Estimated epidemic onset times vs effective distance plot for various diseases. In figure a., the time to first case is calculated using a metapopulation model simulation for a hypothetical disease, assuming infection seeding in Greenville County. In figures b., c., and d., the epidemic onset dates are estimated from hospitalization data. In all figures, the effective distance is calculated using the radiation model.

### Relative timing of epidemic onset correlates with effective distance across seasons

To assess the spatial coherence of epidemic spread, we examined the time to epidemic onset across counties, defined as the difference between each county’s onset date and the earliest onset date observed within South Carolina during a given season. We found that this relative timing was positively correlated with effective distance for influenza across multiple seasons. Specifically, during the 2019–2020 influenza season, the earliest estimated onset occurred in Greenville County, and the correlation between effective distance and time to onset was ρ = 0.52, p<0.001 (Figure 3). In contrast, for the 2022–2023 influenza season, Charleston County exhibited the earliest onset, yet a similar correlation was observed (ρ = 0.66, p<0.001; Figure 3). Importantly, these relationships are held across both types of mobility data (Figure S3). For RSV, we also observed positive correlations between effective distance and onset timing, though the strength of the association was lower during the 2019–2020 season, likely due to limited data availability across counties (Figure S3).

**Figure 3.**
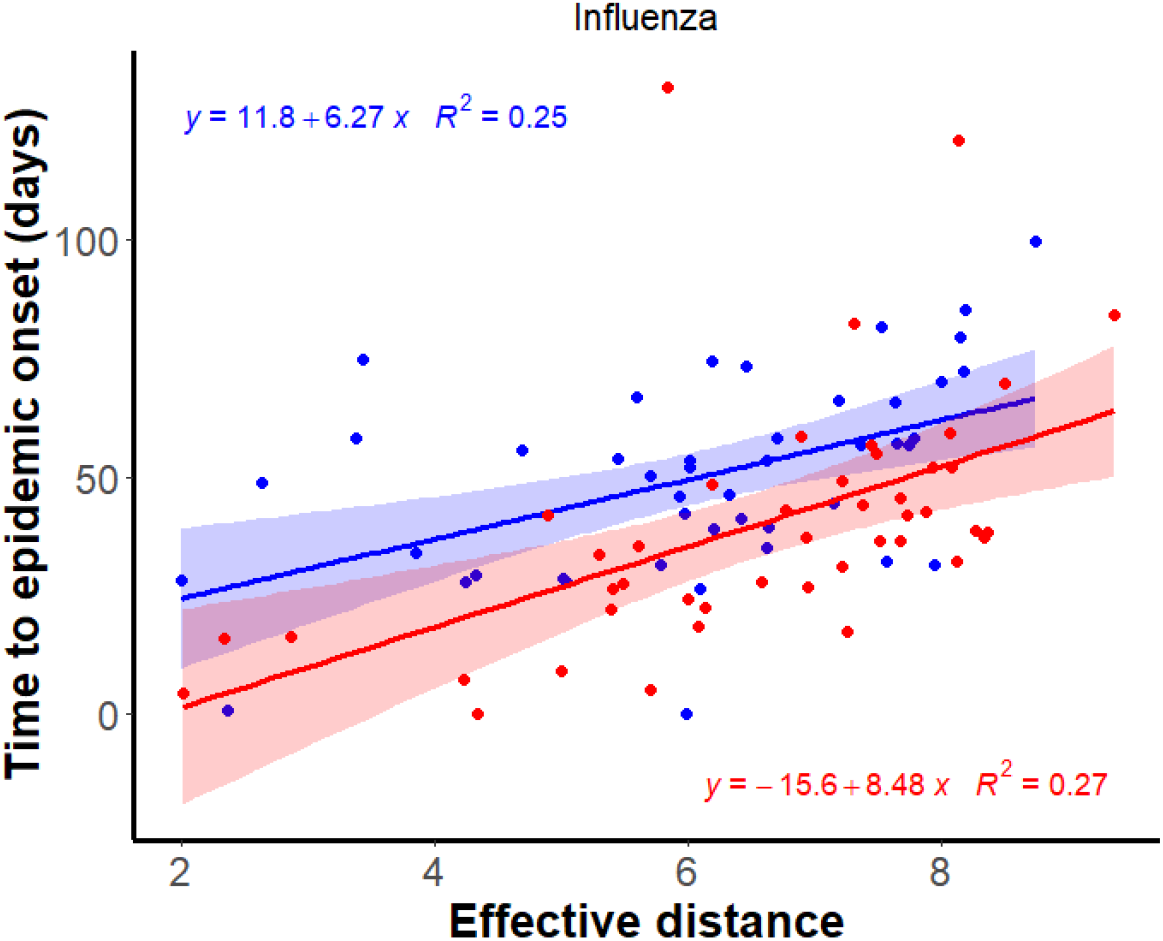
Effective distance is good in predicting epidemic onset times across years for influenza. The blue dots represent time to epidemic onset in different counties, assuming the first epidemic onset in Greenville County for the 2019/2020 season, whereas red dots represent estimates for the 2022/2023 season, assuming the first epidemic onset in Charleston County.

### Effective distance predicts state-level COVID-19 arrival time and Influenza epidemic onset date

At the national scale, we evaluated whether effective distance derived from air traffic networks could predict the spatial progression of respiratory epidemics across U.S. states. Using 2019 air traffic data among major airports, we found a positive correlation between effective distance from Washington state, which is assumed as the initial seeding location and where the first infection was reported, and the timing of the first reported COVID-19 case in each state during the first wave of the pandemic (Spearman’s ρ = 0.63, p<0.001, Figure 4a). This suggests that major air travel routes contributed to the early geographic spread of SARS-CoV-2. We applied a similar analysis to the 2024–2025 influenza season, using 2024 air traffic volume data. Again, effective distance from the earliest-affected state (estimated as Texas) was positively correlated with estimated epidemic onset dates at the state level (ρ = 0.48, p<0.001, Figure 4b).

**Figure 4.**
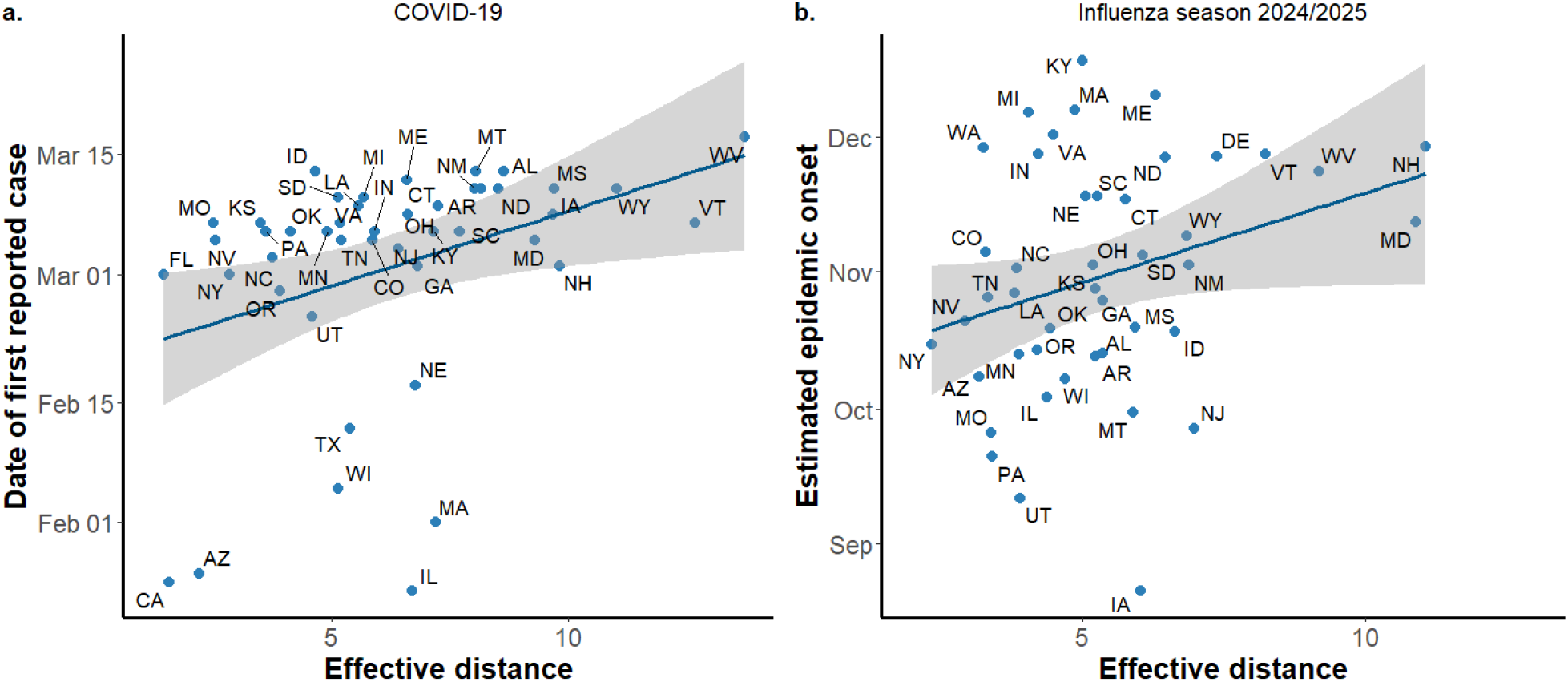
Correlation between effective distance derived from air traffic flow and starting points for COVID-19 and influenza. The left panel shows correlation with the date of the first reported case in each state, and the effective distance is calculated from Washington, where the first infection was reported. The right panel shows estimated epidemic onset times for the influenza 2024/2025 season, and the effective distance is calculated from Texas, which had the earliest estimated onset date.

## Discussion

Respiratory infectious diseases continue to impose substantial burdens on healthcare systems across the United States, with COVID-19, seasonal influenza, and respiratory syncytial virus (RSV) representing three of the most impactful and recurrent threats [25]. In this study, we developed a simple metapopulation modeling framework that leverages both empirical data and a theoretical mobility model to estimate the relative timing of epidemic onset for these pathogens. We demonstrate the flexibility and scalability of our model by predicting county-level epidemic progression within South Carolina and state-level introductions across the United States. These predictions offer a valuable decision-support tool for guiding the proactive allocation of healthcare resources and personnel, both in the context of emergent epidemics and in anticipation of recurrent seasonal outbreaks. The ability to generate spatial forecasts without requiring detailed epidemiological parameters makes this approach particularly useful for early-phase response planning when data is limited and time-sensitive decisions are critical.

Our findings highlight the potential utility of mobility-informed models as tools for strategic public health decision-making, particularly in forecasting the early geographic trajectory of infectious diseases [7,11,14,15,41]. Consistent with established patterns observed in previous pandemics and seasonal epidemics, we find that infections tend to emerge first in regions with high population density and extensive connectivity, typically large urban counties or states with major transportation hubs, before diffusing outward to smaller, less connected areas [42–45]. This directional flow from population centers to peripheral regions was consistent across multiple respiratory pathogens and across different seasons, suggesting a generalized mechanism of spatial disease spread. Notably, even when initial seeding events occur in remote or sparsely populated counties, infections are more likely to propagate toward urban centers, which subsequently act as secondary hubs, amplifying transmission and facilitating further spatial diffusion. The robustness of these spatial patterns across disease systems underscores the value of such models for anticipating the spread of novel pathogens. Even in the absence of precise epidemiological parameters, such as transmission rates or incubation periods, mobility-informed frameworks can generate actionable insights into the likely progression of an emerging outbreak, enabling earlier interventions and more targeted deployment of limited public health resources [7,41].

The success of simple analytical and statistical mobility models, such as gravity and radiation models, in capturing large-scale human mobility patterns underscores a fundamental degree of regularity and predictability in human movement [33,46–49]. This predictability, in turn, can be harnessed to model the spatial diffusion of infectious diseases, particularly during the early stages of an outbreak when real-time epidemiological data may be sparse. Prior work has demonstrated the efficacy of mobility-informed models for predictions. For instance, Brockmann and Helbing (2013) employed global air traffic data to reconstruct the spatial progression of the 2001 SARS outbreak and the 2009 H1N1 influenza pandemic using effective distance [7]. In our study, although we primarily focus on epidemic onset dates rather than the arrival times, we similarly find that the effective distance enables accurate prediction of the relative timing of disease arrival at both county and state levels. These results align with recent findings by Babazadeh Maghsoodlo et al. (2025), who reported a strong linear correlation between effective distance and “overtaking time,” defined as the point at which within-location/patch dynamics dominate over spatial seeding (between-patch dynamics) [41]. Their study, consistent with ours, suggests that effective distance is not only predictive of epidemic onset, but may also serve as a robust proxy for other epidemic phases. Together, these findings support the broader applicability of effective distance as a unifying framework for spatio-temporal epidemic forecasting.

Variation in the predictive accuracy of our model across disease systems, COVID-19, influenza, and RSV, highlights the importance of epidemiological context and transmission dynamics. For instance, while the early phase of COVID-19 in South Carolina exhibited relatively strong agreement with model predictions, greater variability was observed in estimated arrival times at the national scale. This likely reflects the reality of multiple, near-simultaneous introductions of SARS-CoV-2 into the United States through major international air travel hubs, including California, Illinois, and New York [50]. The presence of multiple seeding events undermines the assumption of a single-source introduction, resulting in spatial decoherence that reduces model accuracy. Similar dynamics can occur in the context of seasonal respiratory diseases such as influenza and RSV [51]. Even at the county level, concurrent epidemic onset in multiple densely populated counties driven by local transmission or residual endemic circulation can generate overlapping waves of infection that complicate efforts to infer a clear directional spread. In such scenarios, disease resurgence during the typical influenza season may not require new external introductions but may instead result from the amplification of existing, low-level transmission chains. Nonetheless, if surveillance systems can identify and distinguish newly emergent strains, such as novel influenza subtypes, our model is likely to perform with greater accuracy, as strain-specific spread would resemble a more classic invasion process with well-defined geographic origins. For example, a recent genomic analysis of influenza virus sequences collected during 2014 to 2023 suggests a combination of commuting patterns and lineage competition to be a strong predictor of seasonal influenza epidemics in the US [52].

While our modeling framework offers a flexible and data-efficient approach to predicting spatial patterns of disease spread, several limitations warrant consideration. First, the mobility data used in this study, specifically county-level commuting flows and annual airline traffic volumes, are inherently static and may not capture temporal fluctuations in population movement, such as those driven by school calendars, holidays, or behavioral changes during outbreaks. Incorporating more dynamic, high-resolution mobility datasets (e.g., real-time mobile phone data or weekly traffic patterns) could substantially enhance the model’s predictive accuracy, particularly during rapidly evolving epidemic conditions. Second, our analyses rely on retrospectively estimated epidemic onset dates, which, although suitable for model validation, do not reflect real-time decision-making contexts. In practice, prospective forecasting would require robust, near-real-time surveillance systems capable of detecting local epidemic activity promptly. This is particularly critical in high-risk locations, those with high population density or mobility centrality, where early signals of outbreak emergence are most likely to appear. Finally, while our model is based on a simple SIR framework, it could be extended to incorporate additional epidemiological complexities, including age-structured transmission, vaccination dynamics, hospitalization, and mortality. Such extensions would improve the model’s utility not only for spatial prediction but also for estimating healthcare burden and evaluating intervention strategies.

Our study demonstrates that metapopulation models informed by human mobility, whether derived from empirical commuting and air traffic data or theoretical frameworks like the radiation model, can provide robust predictions of the relative timing of respiratory disease spread across spatial scales. By leveraging the structural regularities in human movement, we show that effective distance serves as a powerful, low-data metric for anticipating the geographic progression of pathogens such as COVID-19, influenza, and RSV. The consistency of spatiotemporal patterns across pathogens and seasons underscores the utility of mobility-informed modeling for early-phase outbreak response and strategic planning. Importantly, these models remain valuable even in the absence of detailed epidemiological parameters, making them especially useful in real-time decision-making contexts. As global connectivity and pathogen emergence continue to rise, incorporating scalable, mobility-based models into public health infrastructure, alongside enhanced surveillance systems, offers a promising pathway for improving epidemic preparedness and response.

## Data Availability

All data produced in the present study are available upon reasonable request to the authors.

