## Supplemental Figures for "Mobility-informed metapopulation models predict the spatio-temporal spread of respiratory epidemics across scales"

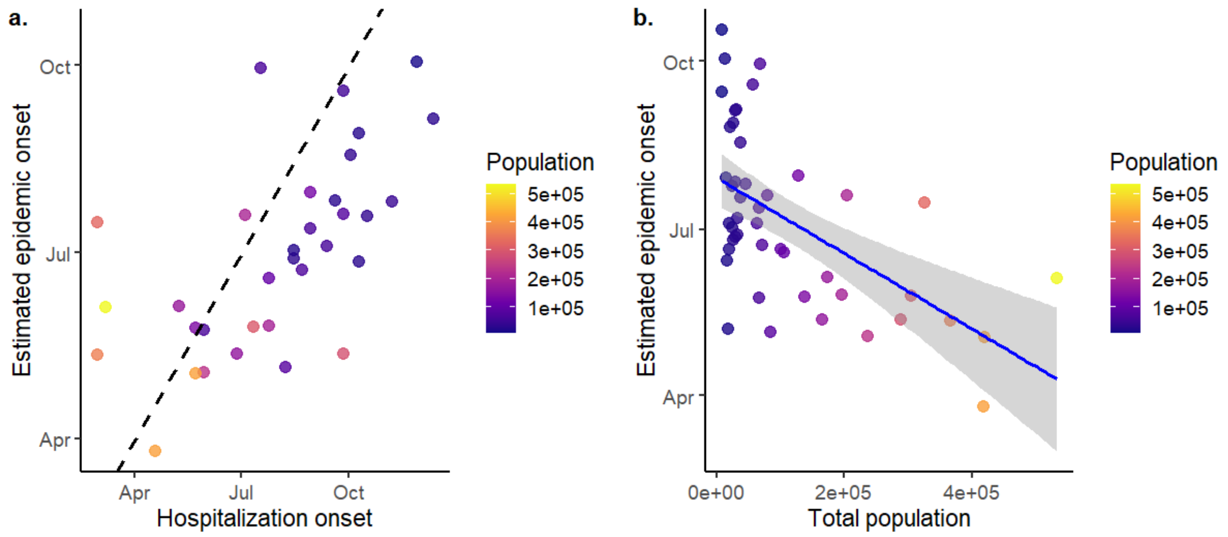


Figure S1. Estimated epidemic onset dates for the RSV 2022/2023 season.

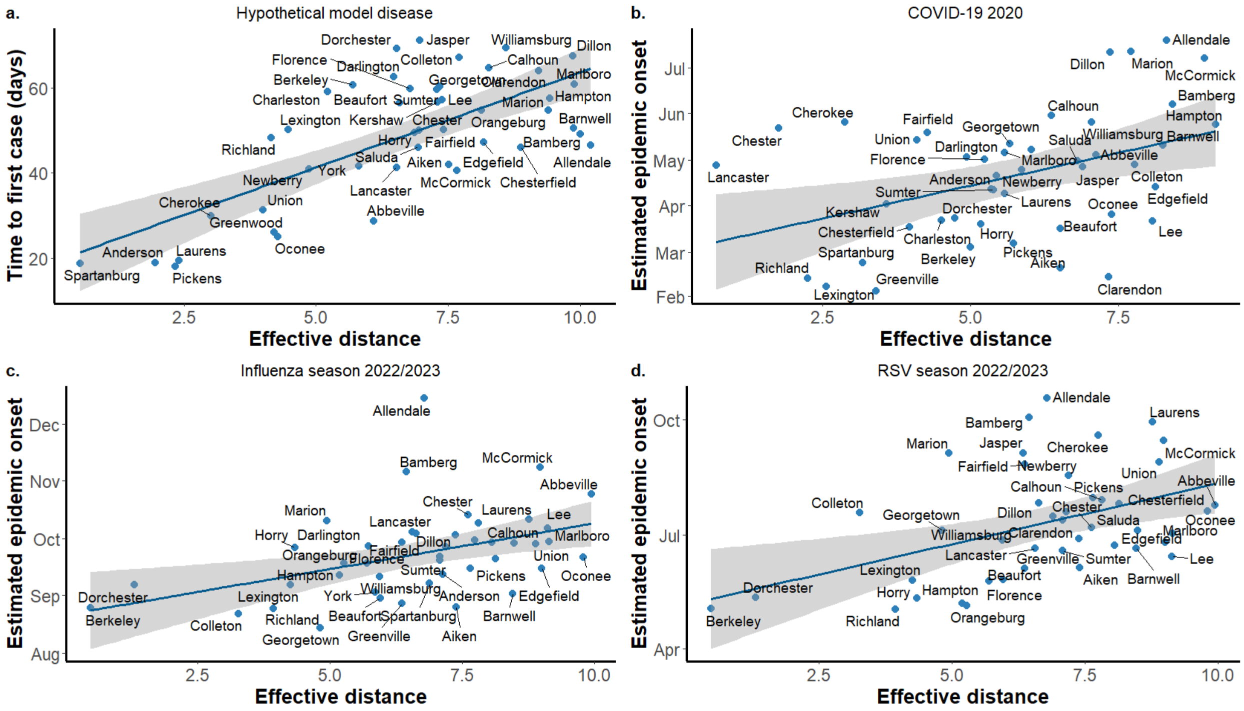


Figure S2. Estimated epidemic onset times vs effective distance plot for various diseases. In figure a., the time to first case is calculated using a metapopulation model simulation for a hypothetical disease, assuming infection seeding in Greenville County. In figures b., c., and d., the epidemic onset dates are estimated from hospitalization data. In all figures, the effective distance is calculated using the county-level commuting data.

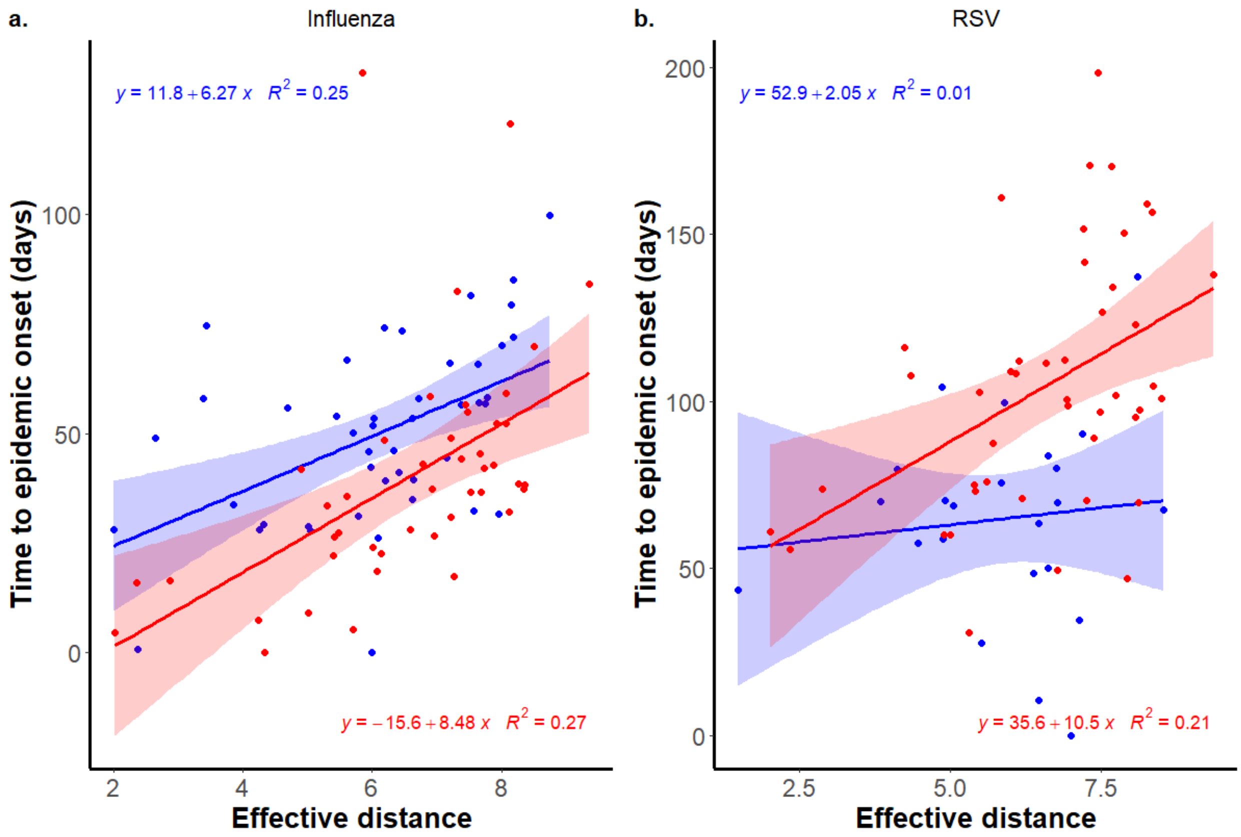


Figure S3. Effective distance vs time to epidemic onset for influenza (left) and RSV (right) across two seasons. The blue dots represent time to epidemic onset in different counties, assuming the first epidemic onset in Greenville County for the 2019/2020 season, whereas red dots represent estimates for the 2022/2023 season, assuming the first epidemic onset in Charleston County.
